# Development and Initial Evaluation of a Patient Decision Aid to Support Decision-Making in Care-seeking Patients with Subacromial Pain Syndrome in Primary Care

**DOI:** 10.1101/2025.01.17.25320492

**Authors:** Kristian D Lyng, Samantha C Bengtsen, Joshua R Zadro, Nadine E Foster, Jens L Olesen, Janus L Thomsen, Jens Søndergaard, Glyn Elwyn, Rhiannon E. Evans, Peter Malliaras, Véronique Lowry, François Desmeules, Michael S Rathleff

**Affiliations:** Center for General Practice, Aalborg University, Aalborg, Denmark; Department of Health Science and Technology, Aalborg University, Aalborg, Denmark; Sydney Musculoskeletal Health, The University of Sydney, Sydney, New Souths Wales. Australia; Institute for Musculoskeletal Health, Sydney Local Health District, Sydney, New South Wales, Australia; STARS Education and Research Alliance, Surgical Treatment And Rehabilitation Service (STARS), The University of Queensland and Metro North Health, Brisbane, Australia; Research Unit of General Practice, Department of Public Health, University of Southern Denmark, Odense, Denmark; The Dartmouth Institute for Health Policy and Clinical Practice, Lebanon, NH, USA; Centre for Development, Evaluation, Complexity and Implementation in Public Health Improvement (DECIPHer), Cardiff University, Cardiff, UK; Department of Physiotherapy, School of Primary and Allied Health Care, Monash University, Melbourne, Australia; School of Rehabilitation, Medicine Faculty, University of Montreal, Montreal, QC, Canada; Orthopaedic Clinical Research Unit, Maisonneuve Rosemont Hospital Research Center, Montreal, QC, Canada

**Keywords:** Decision aid, elbow & shoulder, mixed-method study, musculoskeletal disorders, pain, primary care, shared decision-making, shoulder, mixed-method study, qualitative research

## Abstract

**Objectives:** To describe the development and initial evaluation of a new context-specific patient decision aid for care-seeking patients with subacromial pain syndrome (SAPS) in primary care.

**Methods:** To develop a new contextually relevant decision aid, mixed methods research study with four components was conducted. We adapted a previously tested decision aid using the ADAPT guidance (Activity I) and simultaneously developed new items using the International Patient Decision Aid Standards and the Ottawa Decision Support Framework (Activity II) to inform a prototype of the decision aid. In activity III, we alpha tested the prototype through think-aloud interviews with 10 patients with SAPS and 10 healthcare practitioners of different disciplines. In the interviews, participants were also asked to rate their readiness for the decision-making process using the Preparation for Decision-Making (PrepDM) questionnaire and the face-validity of the prototype was evaluated using the QQ-10 questionnaire. Revisions were made based on the feedback from the participants, the project group and a reference group consisting of 26 individuals including patients and healthcare practitioners. Following this, the new prototype was beta-tested in primary care with 23 care-seeking patients with SAPS. All participants were asked to complete the Shoulder Pain and Disability Index (SPADI), EQ-5D-3L and Decisional Conflict Scale (DCS) before, after and two weeks after introduction to the decision aid. Participants also scored the Decision Regret Scale (DRS) after two weeks. Following beta-testing, 13 interviews were conducted to explore the acceptability and usability of the decision aid, guided by the Standards for UNiversal reporting of Decision Aid Evaluations (SUNDAE) guidelines and Preparation for Decision Making Scale (PrepDM).

**Results:** Based on the two first activities, the prototype decision aid included 10 treatments. Findings from alpha-testing highlighted that the prototype was acceptable and useful in preparing both patients and healthcare practitioners for the treatment decision-making process. Beta-testing showed that after introduction to the decision aid, the DCS decreased from 40 ± 18 at baseline to 25 ± 18 two weeks after the decision, indicating low levels of decisional conflict. Furthermore, after two weeks the DRS indicated low levels of decisional regret (25 ± 9). SPADI and EQ-5D-3L scores were largely similarly across all time-points. Interviews highlighted that both patients and healthcare practitioners felt the decision aid was a valuable tool for clinical practice.

**Conclusions:** Our decision aid is a promising tool for influencing the decision-making process in patients with SAPS in primary care. Further research that compares the offer of the decision aid to patients and healthcare practitioners in primary care with usual care is needed.

**Practice Implications:** Further research is needed to fully evaluate the effects of the decision aid.

**Support and Sponsor:** This study was funded by Novo Nordisk Foundation, TrygFonden, Danish Association of Physiotherapy and Aalborg University. NEF is funded through an Australian National Health and Medical Research Council (NHMRC) Investigator Grant (ID: 2018182). JRZ is funded through an Australian National Health and Medical Research Council (NHMRC) Investigator Grant (ID: APP1194105). None of the funders were involved in the research. The sponsor is non-commercial and declares no conflicts of interest.

## Background

Shoulder pain poses a significant societal burden where 1 to 5% of patients from primary care experience shoulder pain, with a 26% increase in incidence rate from 2005-2017 [1,2]. Subacromial pain syndrome (SAPS), often also called rotator cuff-related shoulder pain, is the most reported condition causing shoulder pain [1,3]. The long-term prognosis can be poor with some studies reporting a 12-month recovery from 32 to 59% [4–6]. Clinical practice guidelines recommend simple analgesics (i.e., acetaminophen) as a first-line pharmacological treatment in the acute stage, often accompanied by conservative treatments such as exercise [7,8]. Other strategies include manual therapy, injection therapies, and surgery may be introduced in later stages [9]. Recent evidence has highlighted the limited effectiveness of current treatments, leading healthcare practitioners to offer patients alternative and potentially less beneficial options, such as shockwave therapy, platelet-rich plasma injections, or acupuncture [7,9–11]. Clinically, this suggests that decision-making related to the management of SAPS is a preference-sensitive process, requiring careful deliberation of the individual attributes of existing treatments, the patients’ needs and their values [12].

Previously, we assessed the needs, preferences, and priorities of individuals with shoulder pain, including SAPS. In one study, we explored the decisional needs of patients living with SAPS, emphasizing the complexity of the treatment decision-making process and patients’ potentially unmet decisional needs [13]. We demonstrated that patients preferred to feel involved and supported in the decision-making process, and thus, wanted improved access to adequate information about their condition and treatment options [13]. In our second study, we explored patient preferences by identifying key attributes related to treatment through a systematic review and interviews with stakeholders [14]. The identified attributes (e.g., pain reduction, impact on quality of life) helped us understand the preferences of patients with SAPS, thus allowing a more personalised deliberation of potential uncertainty that arises due to the preference-sensitive process. Lastly, we collected research priorities from over 600 patients with shoulder pain and healthcare practitioners and prioritised them into a top 10 list [15]. These priorities highlighted the need for greater focus on translating existing evidence into clinical practice and improving patient education [15]. One approach to achieve this, is using decision aids, which are developed to support patients in the decision-making process [16]. Decision aids provide information about existing treatment options using different methods, often including visuals and icon arrays [16]. A non-systematic literature search of decision aids to support treatment decision-making in shoulder pain identified a co-designed patient decision aid developed by Zadro et al. [17,18]. This work showed that the decision aid had minimal impact on participants’ treatment intentions, attitudes, informed choice, or decisional conflict, however, it did show a small improvement in knowledge [18]. The authors attributed the lack of effect to several factors including that the decision aid was not delivered by a healthcare professional, leaving no room for tailored guidance and the ability to address any concerns or questions from the patient. Furthermore, the decision aid was introduced online and only when patients were already on a waitlist for surgery, which may pose as unfavourable conditions for influencing decision-making as patients might be too far in the decision-making process [18]. To our knowledge, no decision aids have been specifically developed and tailored to support the decision-making process in care-seeking patients with SAPS in the primary care.

This paper outlines the development, adaptation, and initial evaluation of a decision aid specifically designed to support care-seeking patients with SAPS during consultations with primary care healthcare practitioners.

## Methods

### Study Design

This study was designed as a prospective multi-methods study aiming to develop and evaluate a decision aid. The development process was focused on developing new items, tailoring the previously developed decision aid for use in Danish primary care for care-seeking people with SAPS. To efficiently develop a decision aid for our Danish context, we adapted a pre-existing English-language decision aid following ADAPT and merged it with new treatment strategies to ensure it was sensitive to the context (primary care consulters) [19]. The previous decision aid was co-designed for use by people with SAPS considering surgery [17,18]. To adapt the decision aid to our context (Danish primary care and for patients seeking primary care), the International Patient Decision Aid Standards (IPDAS) and the Ottawa Decision Support Framework (ODSF) were used to develop new items for the decision aid [16,20]. The reporting of the study follows the Standards for UNiversal reporting of Decision Aid Evaluations (SUNDAE) guidelines [21]. The protocol was pre-registered at Open Science Framework [22]. Visualisation of all the study phases and activities from the full procedure can be seen in **Figure 1**. The description of phase 3 and activity V is outside the scope of this paper. The study was conducted in accordance with the Helsinki Declaration and was deemed exempt from full ethical approval by The North Denmark Region Committee on Health Research Ethics (2023-000206).

**FIGURE 1.**
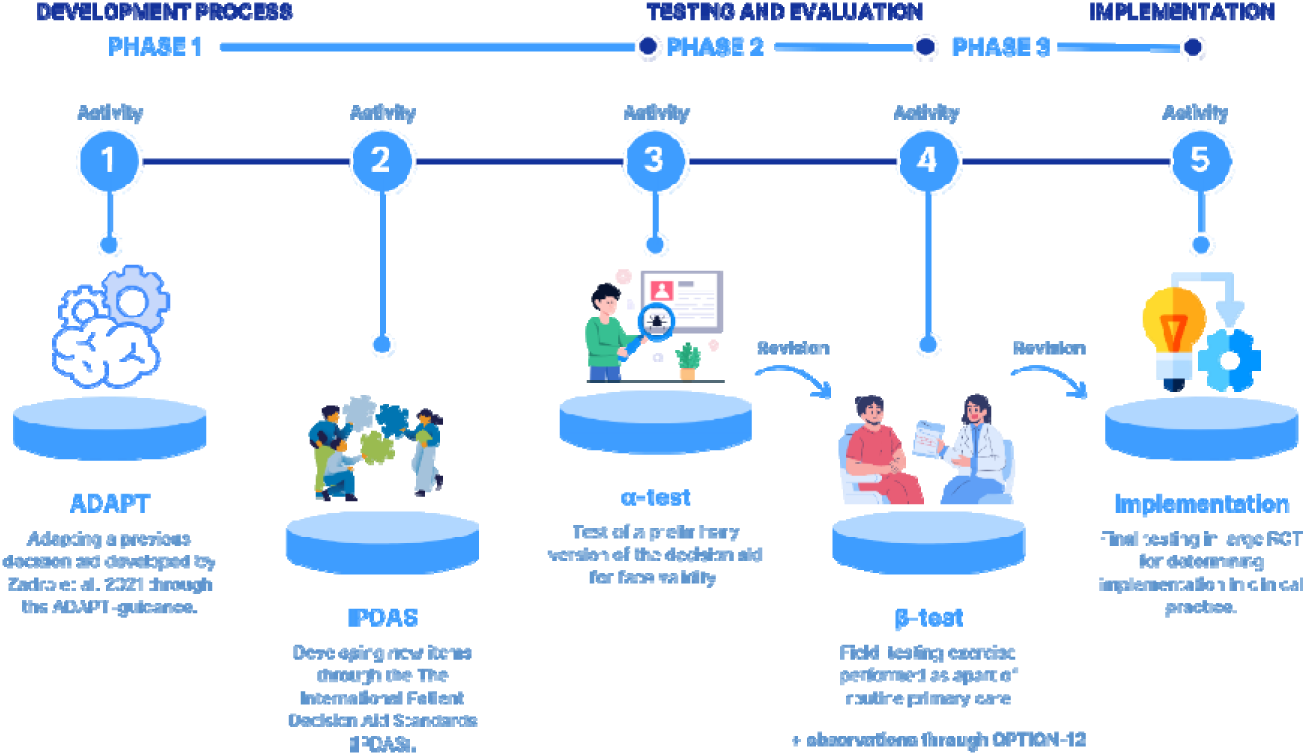
DEVELOPMENT AND EVALUATION PROCESS. Implementation was not part of this study.

### Target group(s) and context

The primary target group for our decision aid is patients diagnosed with non-traumatic SAPS seeking primary care through a first-contact practitioner in Denmark (i.e. general practitioners and physiotherapists). During alpha-testing, adults currently living with or recovered from SAPS were included if they: 1) self-identified as having or had shoulder pain for ≥ 3 months, 2) rated their worst shoulder pain in the last week at least ≥ 3/10 on a Numeric Rating Scale and, 3) reported that their shoulder complaint had a non-traumatic origin. All licensed healthcare practitioners involved in the treatment of SAPS was eligible to participate and provide feedback on the prototype. For beta-testing, adults were included if they were experiencing antero-lateral shoulder pain for a least a week. Furthermore, they needed to have a history of activity-related shoulder pain and an atraumatic onset. Lastly, all participants needed at least three positives on the following five diagnostic tests: Hawkins-Kennedy test, Neer’s test, painful arc, Resisted External Rotation test and Jobe’s test [23]. Healthcare practitioners from primary care (physiotherapists and general practitioners) were included in the study if they treated at least five cases of SAPS per year. The healthcare practitioners were recruited using local professional networks and needed to have no prior knowledge about the decision aid.

### Project and Patient and Public Involvement groups

Two steering groups were established: a project group and a patient and public involvement (PPI) group. The project group consisted of thirteen researchers with different demographics, experience, professional backgrounds, and nationalities. Of these, eight were physiotherapists, four were medical doctors and one was a social scientist. The project group members came from five countries including Australia, Canada, Denmark, United Kingdom, and United States of America. The PPI group consisted of 26 Danish people with various backgrounds (six people living with SAPS, two informal caregivers, three physiotherapists, two general practitioners, two orthopaedic surgeons, a rheumatologist, an occupational medicine doctor, a chiropractor, a nurse, and a psychologist). Of these 26 people, two were also policymakers, two were researchers, and two representatives outside the healthcare system. All participants were recruited from either previous studies, social media, or social networks. Full demographic data has been described elsewhere [24].

### Phase 1: Development process

The study’s first phase aimed to develop the first draft of the new decision aid. This process consisted of two parallel activities (**I** and **II**), including adapting the previous decision aid (I) and developing new items to fit this new context (II). The rationale for adapting the decision aid is described in **Appendix 1**.

### Activity I: Adapting a previous decision aid

In the first activity, the lead author of the paper was invited into the project group and the decision aid was adapted to our Danish context using the ADAPT guidance and its four iterative steps [19] (See figure 2). A detailed description of the ADAPT process is provided in **Appendix 1**, along with a comprehensive overview of the decision aid developed by Zadro et al., structured according to the Template for Intervention Description and Replication (TIDieR) framework [25] (**Appendix 2**). The PPI group agreed on the need for a new decision aid and the suitability of such an aid for clinical practice. A formal translation of the original decision aid was undertaken from English to Danish through the Translation, Review, Adjudication, Pre-testing, Documentation (TRAPD) approach (See **Appendix 3**) [26].

**Figure 2.**
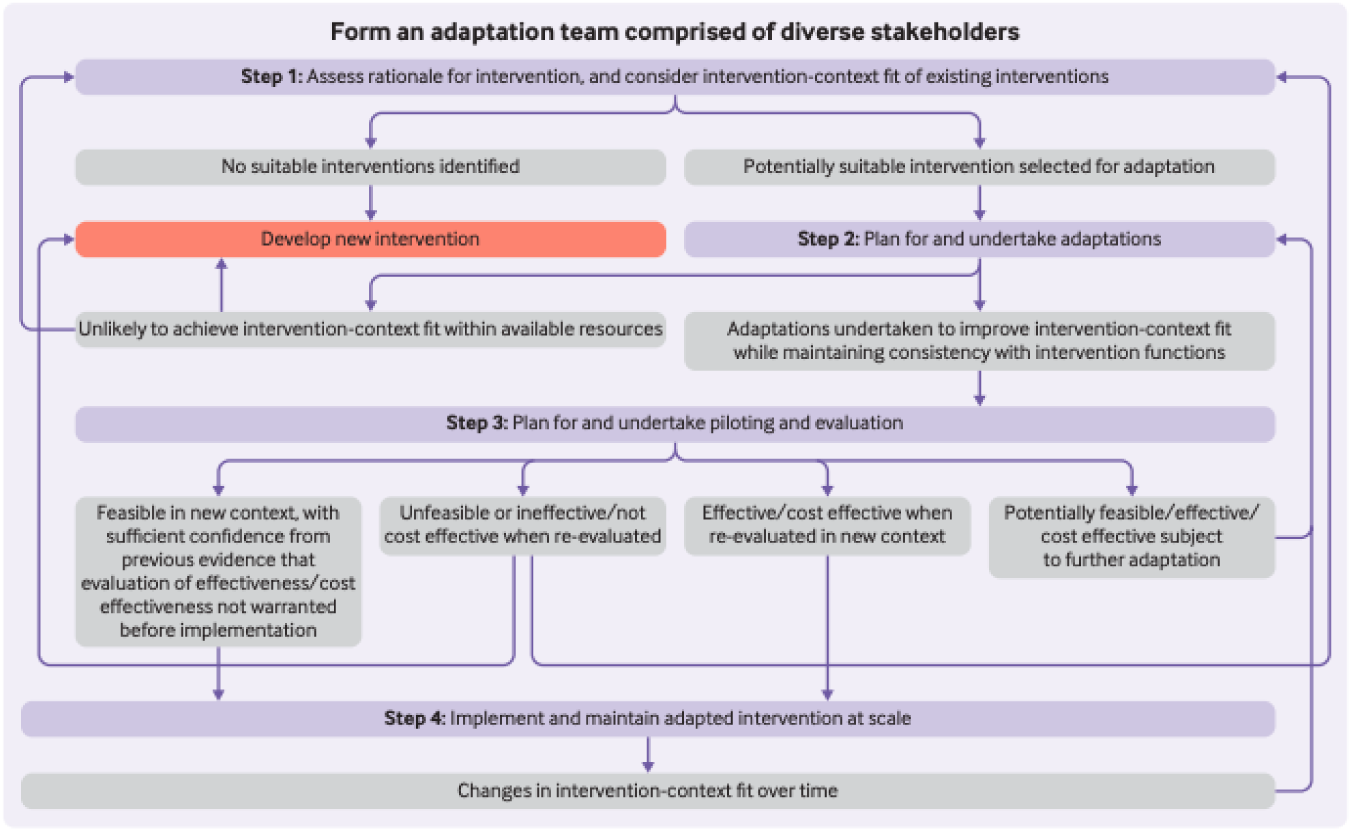
ADAPT PROCESS. Used from Moore et al. Step 4 was not included in this study.

### Activity II: Developing new items

In the second activity, new items for the decision aid were developed using the IPDAS and ODSF. The previous decision aid primarily focused on providing in-depth information about surgical options and offered limited information on non-surgical treatments. Thus, there was a need to expand the content to better cover non-surgical treatment options, which are more relevant for our context. Firstly, we identified the most prevalent treatment options for SAPS through reviewing clinical guidelines, systematic reviews, and reports of common management strategies used in practice [7–10,27–29]. Since there is no data on the most frequently used treatments in Denmark for SAPS, the PPI group considered information about clinical guidelines, results from trials, usual care-pathways, and the content from the original decision aid. The PPI group members were invited to provide inputs on other treatments that needed to be included in a prototype of the new decision aid. Potential suggestions were discussed and included based on consensus between the PPI and project group. Benefits and harms were extracted from relevant studies reporting effects and adverse events of the respective treatments. We prioritised systematic reviews and meta-analyses due to their comprehensive synthesis of evidence, followed by randomised control trials for their robust methodology in evaluating treatment efficacy and safety [30,31]. In cases of limited evidence, high-quality observational studies were considered. In cases of complete lack of knowledge on benefits and harms, this was noted for the users to consider. The extracted data were compiled in icon arrays to visualise the benefits and harms of the respective treatments, including different follow-up timepoints when available. Importantly, we primarily focused on outcomes at the longest duration (e.g., 12 months or more). After the collection of data, a prototype was created inspired by the format used in Zadro et al., the decisional needs gathered previously, and by the IPDAS checklist.

### Phase 2: Testing and evaluation of the decision aid

The second phase consisted of two activities (**III** and **IV**) which were alpha-testing of a prototype of the decision aid (**Activity III**) and beta-testing (**Activity IV**). Alpha-testing serves as an essential step in refining the quality of a decision aid in non-clinical settings and exploring face validity. It focuses on a small, group of healthcare professionals and people living with SAPS to identify and address potential issues in its design and content. This stage allowed for iterative feedback and adjustments before beta testing, which involved a broader, more representative sample of actual users within clinical practice settings with primary care healthcare practitioners and care-seeking patients diagnosed with SAPS.

### Activity III: Alpha-testing

Preparation for alpha-testing began with contacting patients with current or previous SAPS and healthcare practitioners who manage this condition and inviting them to participate. Relevant healthcare practitioners and patients within the target group and context were recruited through professional and social networks. Alpha-testing consisted of think-aloud interviews where the participants read the prototype material out loud [32,33]. While reading the prototype, the participants were invited to share their thoughts about functionality, usability, and content [32]. The interviews were conducted by one author from the project group (SCB) either in person or online via Microsoft Teams. Consent forms and background information from the participants were obtained through the online application REDCap^TM^ [34,35]. The interviews were audio recorded for use in the data analysis. After the interviews, participants were given time to read through the prototype again, before answering the Preparation for Decision-Making (PrepDM) questionnaire and six questions from the QQ-10 questionnaire [36,37]. The QQ-10 questionnaire is designed to evaluate patients’ perspectives (face-validity) on the value and burden associated with using health-related quality of life assessment tools. Two Danish editions of the PrepDM were used, one for patients and one for healthcare practitioners [38,39], alongside a non-validated translated version of the QQ-10. The QQ-10 questions included “*The questionnaire helped me to communicate about my condition*”, “*The questionnaire was relevant to my condition*”, “*The questionnaire was easy to complete*”, “*The questionnaire included all the aspects of my condition that I am concerned about”*, “*The questionnaire was too long*”, “*The questionnaire was too complicated”* and were rated on a 5-point Likert response scale rating from “*Strongly agree*” to “*Strongly disagree*” [37]. Audio recordings of the interviews were anonymised and transcribed by SB. The interviews were reflectively analysed through an inductive thematic analysis to identify potential changes that might improve the decision aid [40,41]. Revisions were then made by KDL and SB, then the decision aid was forwarded to both the project and PPI group for feedback. Changes were implemented based on the feasibility of the change and the importance as judged by the PPI and project group. After final revisions, the decision aid was forwarded to a graphic designer for optimisation of the visual material.

### Activity IV: Beta-testing

Beta-testing was performed to conduct an initial evaluation of the decision aid as a part of clinical practice with patients and general practitioners or physiotherapists. Participants were informed by telephone about the study before their first encounter with their healthcare practitioners and further informed during the clinical assessment prior to enrolment in the research study. Prior to consultation (i.e., baseline), participants completed the Shoulder Pain and Disability Index (SPADI) [42,43], the EQ-5D-3L [44], the Decisional Conflict Scale (DCS) [45]. The SPADI score is a valid and reliable patient-reported outcome measure assessing pain and functional limitations, scored from 0 (best) – 100 (worst) [43,46]. EQ-5D-3L is used to assess quality of life through domains related to mobility, self-care, usual activities, pain/discomfort, and anxiety/depression [44]. The EQ-5D-3L score is on a scale from -0.594 to 1, where a score of 1 represents full health, a score of 0 corresponds to a health state equivalent to being dead, and negative scores indicate a health state considered worse than death [44]. To assess participants’ readiness to engage in the treatment decision-making process, they were asked to state their current stage of decision making [47]. Participants were presented with six statements and were asked to select the one they agreed with most. The statements included “*I haven’t started thinking about the options*”, “*I haven’t started thinking about the options, but I am interested in doing so*”; “*I am currently considering the options*”, “*I am close to making a decision*“, “*I have already made a decision but am still open to reconsidering*” or “*I have already made a decision and am unlikely to change my mind*” [47]. The patients were introduced to the decision aid by their practitioner. The healthcare practitioners were given no formal instructions in how to introduce the decision aid. After the consultation and introduction to the decision aid, participants were asked to complete the SPADI, EQ-5D-5L, and DCS again on an electronic device in the clinic. Two weeks after enrollment, participants were asked again to complete SPADI, EQ-5D-5L, DCS, and in addition the Decisional Regret Scale (DRS) through REDCap [48]. The DRS includes five questions measuring the distress or remorse after a (health care) decision in clinical practice, scored from 0 (no regrets) – 100 (high regret) [48]. Following this data collection, all participants were invited to a telephone interview where they were asked to elaborate on their experience using the decision aid. The interviews followed the SUNDAE checklist to explore the fidelity of the decision aid in terms of which components of the decision aid were used, to what degree it was delivered and used as intended and anticipated and unanticipated consequences [21]. At the end of the interviews, both patients and the healthcare practitioners who introduced the decision aid were asked to rate its acceptability. Separate questions were tailored to both groups and informed by the QQ-10 [37], Ottawa Hospital Research Institute acceptability questionnaire [49] and the PrepDM [36]. See **Appendix 4** for the full interview guide and the acceptability questions for each interview group. Demographics were not collected from healthcare practitioners. The interviews were analysed through a similar approach as the alpha-testing. Descriptive information was reported using mean, standard deviations and range for continuous data and frequency and percentage for categorical data. Changes based on the beta-testing were discussed within the PPI and project groups, and any changes were described using the framework for reporting adaptations and modifications to evidence-based interventions (FRAME) [50].

## Results

### Phase 1: Development process

#### Activity I: Adapting the previous decision aid

The previous decision aid from Zadro et al. was translated to Danish using the TRAPD-method (see full translation in **Appendix 3**). Adjustments were made to the language and word choices as well as the grammar. The PPI group suggested adjustments to wording, so that the lay person might better understand it. After translation, parts of it were included in our decision aid, and parts of it were excluded. The excluded parts of the previous decision aid were the sections about non-surgical options and the section named “Key message”. The sections about non-surgical options were removed because the participants expressed that they were redundant because of the new items identified from activity II and because they did not align with the requirements of comprehensively displaying the different treatment options. Furthermore, the think-aloud testing showed that the sections were considered as confusing and interrupted the reading flow, especially the key messages.

#### Activity II: Developing new items

Through activity II, the following treatment options were incorporated: best practice advice, corticosteroid injection, and medication in addition to the exercise and surgical option. Based on the recommendations from the IPDAS checklist and inputs from the PPI group, other people diagnosed with SAPS and the project group, we decided to include the following treatment options: wait-and-see, manual therapy, acupuncture, shockwave, and laser therapy. Wait-and-see was recommended by the IPDAS checklist, and the remaining options were chosen because the healthcare practitioners reported that they were commonly used in their clinical practice. The five treatment options contained information about delivery and effectiveness according to the IPDAS criteria. The prototype created prior to phase 2, can be seen in **Appendix 5**.

### Phase 2: Testing and evaluation of the decision aid

#### Activity III: Alpha-testing

Through the alpha-testing, 20 participants (9 male, 11 females) were interviewed. Of these, ten were patients and ten were healthcare practitioners (two general practitioners, two orthopaedic surgeons and six physiotherapists) (see demographics for **activity III** in **table 1**). None of the participants had experience in the development or evaluation of decision aids. Interviews were conducted by SB and lasted between 25 and 70 minutes. Changes made to the prototype based on alpha-testing consisted of changes to language and wording but also changes to the structure. The different sections of the prototype were combined so that information about each treatment option could be accessed simultaneously. These adjustments were informed by iterative rounds of feedback during the think-aloud process, allowing for continuous refinement to address user preferences and enhance clarity. To transit from a prototype into a physical decision aid, we utilised loose-leaf cards with the different treatment options containing an explanation of the treatment, practical advantages, disadvantages, and considerations on the front page, and benefits and harms on the back page. The six treatment cards were enclosed within a cover that featured a front and back page. The back page listed the project group members, their affiliations, practical information about the decision aid, a link to the online version, the project’s funding sources, and the terms and conditions for using the decision aid. Inside the cover, a detailed description of the intended users was provided, along with instructions for how to use the decision aid. Additional content included an anatomical model of the shoulder, a concise summary of the treatments presented, and a set of questions designed to help patients prepare for their consultations with their healthcare practitioners. Based on the QQ-10 questionnaires, the decision aid was regarded by both patients and healthcare practitioners as an acceptable resource for supporting patient decision making in clinical practice. Based on the PrepDM, the decision aid was regarded as useful in preparing patients for treatment decisions **Table 1**. Examples of the product after activity III, following refinement by the graphic designer, is presented in **Figure 3**. The decision aid created prior to the beta-testing can be seen in **Appendix 6**.

**Figure 3.** VISUAL EXAMPLE OF THE DECISION AID. Removed because of lack of valid translation.

**Table 1.** Participant Characteristics, alpha test.

| Table 1. Participant characteristics – α-test | | | | | | |
| --- | --- | --- | --- | --- | --- | --- |
| Characteristics | N (%) or mean ± SD (range) | | | | | |
| **Number (n)** | 20 |  |  |  |  |  |
| - Healthcare practitioners n,% | 10 (50%) |  |  |  |  |  |
| - Patients n,% | 10 (50%) |  |  |  |  |  |
| **Age, years** | 41 ± 10 (28-64) | | | | | |
| **Sex, female (n, %)** | 9, 45.0% |  |  |  |  |  |
| **Pain duration, months** | 30 ± 44 (4-96) | | | | | |
| **Professional experience, years** | 9.3 ± 4.9 (2-16) | | | | | |
| **PrepDM (Score 1-100)** |  |  |  |  |  |  |
| - Healthcare practitioners | 74 ± 19 (28-91) | | | | | |
| - Patients | 84 ± 13 (55-100.0) | | | | | |
| **QQ-10 (6 questions\*)** | **Q1** | **Q2** | **Q3** | **Q4** | **Q5** | **Q6** |
| - Healthcare practitioners n,% |  |  |  |  |  |  |
| - *Strongly agree* | 2 (20%) | 4 (40%) | 0 (0%) | 2 (20%) | 2 (20%) | 0 (0%) |
| - *Mostly agree* | 6 (60%) | 4 (40%) | 5 (50%) | 4 (40%) | 4 (40%) | 4 (40%) |
| - *Neither agree or disagree* | 2 (20%) | 1 (10%) | 2 (10%) | 2 (20%) | 2 (20%) | 3 (30%) |
| - *Mostly disagree* | 0 (0%) | 0 (0%) | 2 (20%) | 2 (20%) | 2 (20%) | 3 (30%) |
| - *Strongly disagree* | 0 (0%) | 1 (10%) | 0 (0%) | 0 (0%) | 0 (0%) | 0 (0%) |
| - Patients n,% |  |  |  |  |  |  |
| - *Strongly agree* | 3 (30%) | 7 (70%) | 4 (40%) | 2 (20%) | 2 (20%) | 0 (0%) |
| - *Mostly agree* | 5 (50%) | 1 (10%) | 2 (20%) | 5 (50%) | 2 (20%) | 2 (20%) |
| - *Neither agree or disagree* | 1 (10%) | 1 (10%) | 2 (20%) | 1 (10%) | 2 (20%) | 3 (30%) |
| - *Mostly disagree* | 0 (0%) | 0 (0%) | 1 (10%) | 1 (10%) | 3 (30%) | 4 (40%) |
| - *Strongly disagree* | 1 (10%) | 1 (10%) | 1 (10%) | 1 (10%) | 1 (10%) | 1 (10%) |
|  | 0 (0%) | 0 (0%) | 0 (0%) | 0 (0%) | 0 (0%) | 0 (0%) |
| *Was the decision aid: **Q1** = Helpful in communicating my decision; **Q2** = Relevant for my condition; **Q3** = Easy to use; |  |  |  |  |  |  |
| **Q4** = Comprehensive; **Q5** = Too long; **Q6** = Overcomplicated. |  |  |  |  |  |  |

#### Activity IV: Beta-testing

Twenty-five were invited and twenty-three patients were recruited and enrolled from four different clinical practices within North Jutland Region (two physiotherapy clinics (n = 14) and two general practices (n = 9) (See **Table 2** for demographics). The two participants who declined the invitation, was due to illness. Participants were recruited between January 2024 and September 2024. All participants completed all forms throughout the study period. At baseline, the average SPADI score, 31.2 (SD: 18.5; range 6.2-70.8), reflected a low level of disability, while the average EQ-5D-3L score, 0.768 (SD: 0.109; range 0.501-0.86) indicated a moderately good health state at baseline. Baseline DCS, 40.0 (SD: 17.9; range 6.3-81.3), indicated medium to low levels of decisional conflict. Following the introduction of the decision aid, the mean DCS score decreased, and further declined after two weeks, reflecting low levels of decisional conflict (**Table 2**). Two weeks after the initial consultation, the mean DRS indicated low levels of decisional regret. None of the participants had experience in the development or evaluation of decision aids. Of the 23 participants included, 12 patients agreed to participate in an interview about their experience with the decision aid. Furthermore, three out of five healthcare practitioners (two physiotherapists, one general practitioner) who recruited participants also agreed to participate in an interview about their experiences introducing and using the decision aid.

**Table 2.**
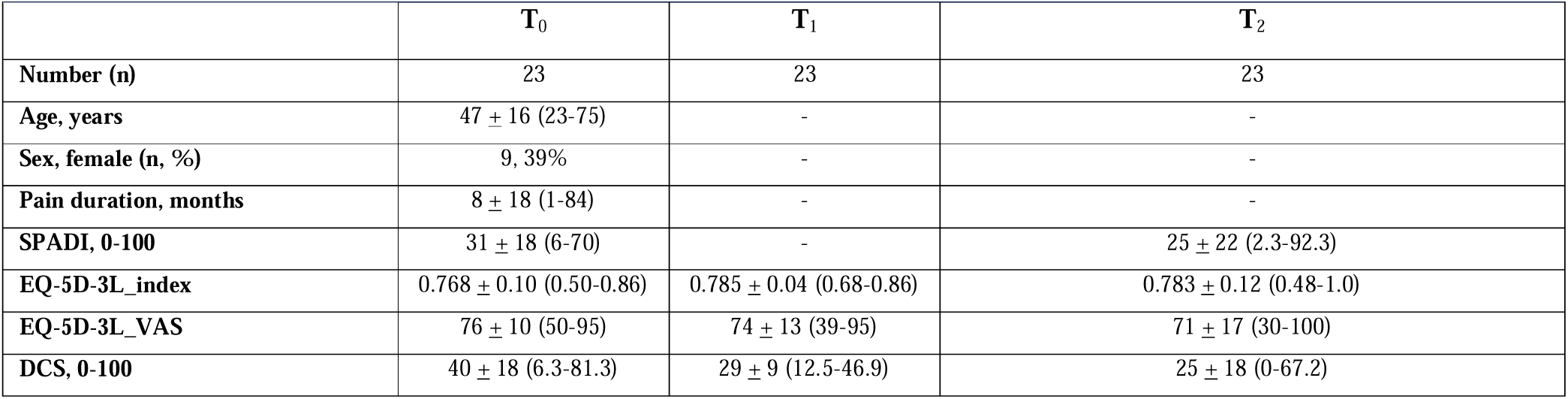
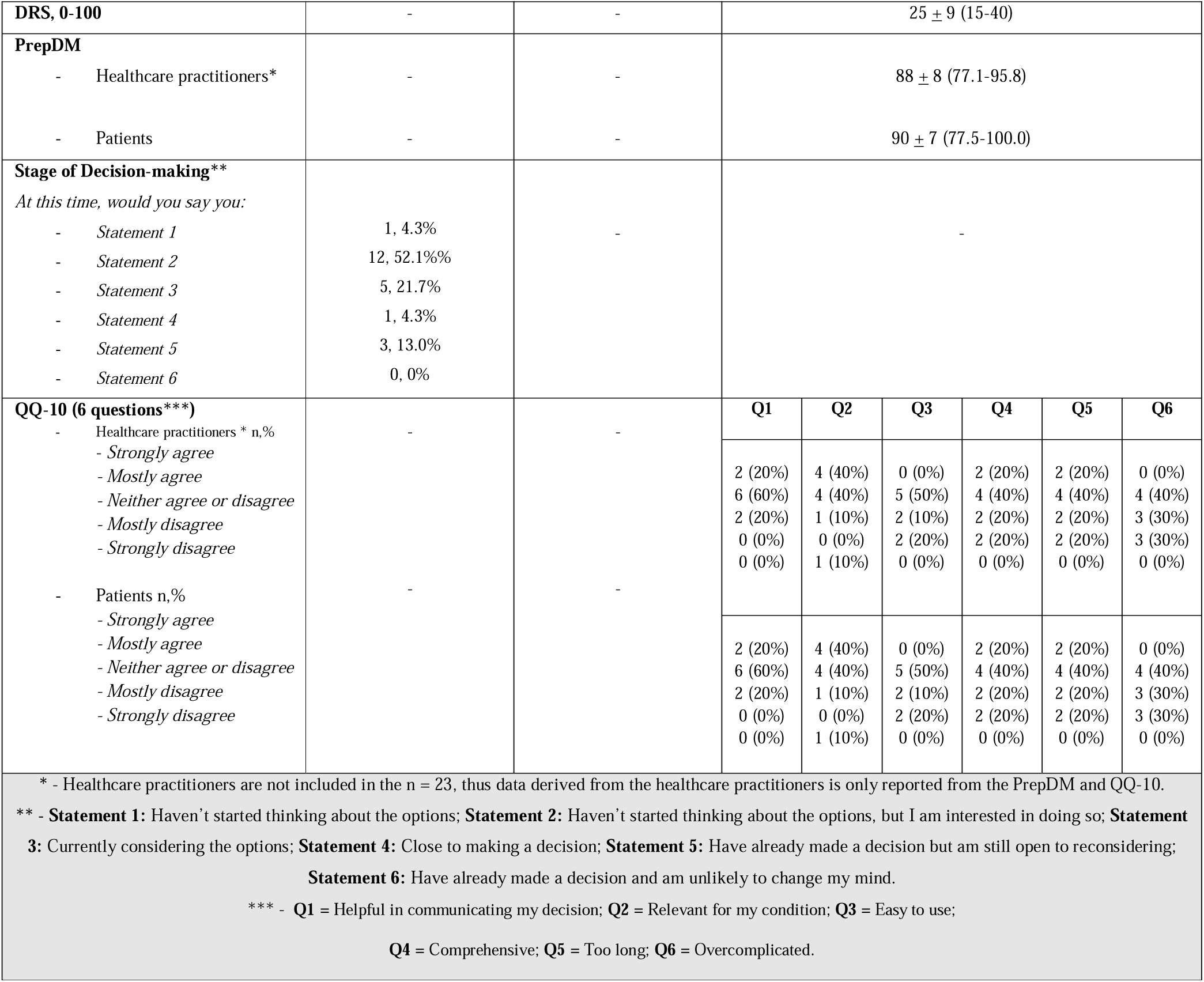
Participants characteristics – Beta-test.

| Table 2. Participant characteristics – β-test | | | |
| --- | --- | --- | --- |
|  | **T<sub>0</sub>** | **T<sub>1</sub>** | **T<sub>2</sub>** |
| **Number (n)** | 23 | 23 | 23 |
| **Age, years** | 47 ± 16 (23-75) | - | - |
| **Sex, female (n, %)** | 9, 39% | - | - |
| **Pain duration, months** | 8 ± 18 (1-84) | - | - |
| **SPADI, 0-100** | 31 ± 18 (6-70) | - | 25 ± 22 (2.3-92.3) |
| **EQ-5D-3L_index** | 0.768 ± 0.10 (0.50-0.86) | 0.785 ± 0.04 (0.68-0.86) | 0.783 ± 0.12 (0.48-1.0) |
| **EQ-5D-3L_VAS** | 76 ± 10 (50-95) | 74 ± 13 (39-95) | 71 ± 17 (30-100) |
| **DCS, 0-100** | 40 ± 18 (6.3-81.3) | 29 ± 9 (12.5-46.9) | 25 ± 18 (0-67.2) |

|  |  |  |  |  |  |  |  |  |
| --- | --- | --- | --- | --- | --- | --- | --- | --- |
| **DRS, 0-100** | - | - | 25 ± 9 (15-40) |  |  |  |  |  |
| **PrepDM** |  |  |  |  |  |  |  |  |
| - Healthcare practitioners* | - | - | 88 ± 8 (77.1-95.8) |  |  |  |  |  |
| - Patients | - | - | 90 ± 7 (77.5-100.0) |  |  |  |  |  |
| **Stage of Decision-making\*\***<br>*At this time, would you say you:* |  |  |  |  |  |  |  |  |
| - Statement 1 | 1, 4.3% | - | - |  |  |  |  |  |
| - Statement 2 | 12, 52.1%% |  |  |  |  |  |  |  |
| - Statement 3 | 5, 21.7% |  |  |  |  |  |  |  |
| - Statement 4 | 1, 4.3% |  |  |  |  |  |  |  |
| - Statement 5 | 3, 13.0% |  |  |  |  |  |  |  |
| - Statement 6 | 0, 0% |  |  |  |  |  |  |  |
| **QQ-10 (6 questions\*\*\*)** |  |  | **Q1** | **Q2** | **Q3** | **Q4** | **Q5** | **Q6** |
| - Healthcare practitioners * n,% | - | - |  |  |  |  |  |  |
| - Strongly agree |  |  | 2 (20%) | 4 (40%) | 0 (0%) | 2 (20%) | 2 (20%) | 0 (0%) |
| - Mostly agree |  |  | 6 (60%) | 4 (40%) | 5 (50%) | 4 (40%) | 4 (40%) | 4 (40%) |
| - Neither agree or disagree |  |  | 2 (20%) | 1 (10%) | 2 (10%) | 2 (20%) | 2 (20%) | 3 (30%) |
| - Mostly disagree |  |  | 0 (0%) | 0 (0%) | 2 (20%) | 2 (20%) | 2 (20%) | 3 (30%) |
| - Strongly disagree |  |  | 0 (0%) | 1 (10%) | 0 (0%) | 0 (0%) | 0 (0%) | 0 (0%) |
| - Patients n,% | - | - |  |  |  |  |  |  |
| - Strongly agree |  |  | 2 (20%) | 4 (40%) | 0 (0%) | 2 (20%) | 2 (20%) | 0 (0%) |
| - Mostly agree |  |  | 6 (60%) | 4 (40%) | 5 (50%) | 4 (40%) | 4 (40%) | 4 (40%) |
| - Neither agree or disagree |  |  | 2 (20%) | 1 (10%) | 2 (10%) | 2 (20%) | 2 (20%) | 3 (30%) |
| - Mostly disagree |  |  | 0 (0%) | 0 (0%) | 2 (20%) | 2 (20%) | 2 (20%) | 3 (30%) |
| - Strongly disagree |  |  | 0 (0%) | 1 (10%) | 0 (0%) | 0 (0%) | 0 (0%) | 0 (0%) |
| * - Healthcare practitioners are not included in the n = 23, thus data derived from the healthcare practitioners is only reported from the PrepDM and QQ-10.<br>** - **Statement 1:** Haven't started thinking about the options; **Statement 2:** Haven't started thinking about the options, but I am interested in doing so; **Statement 3:** Currently considering the options; **Statement 4:** Close to making a decision; **Statement 5:** Have already made a decision but am still open to reconsidering; **Statement 6:** Have already made a decision and am unlikely to change my mind.<br>*** - **Q1** = Helpful in communicating my decision; **Q2** = Relevant for my condition; **Q3** = Easy to use;<br>**Q4** = Comprehensive; **Q5** = Too long; **Q6** = Overcomplicated. |  |  |  |  |  |  |  |  |

### How much of the decision aid and which components were used?

Overall, participants expressed satisfaction with the content of the decision aid, and the visuals were highly praised. Generally, patients expressed that the decision aid contained a lot of different information, however they also stated that the clear structure and appealing visuals made it easy to understand. The colours and the visuals for the icon-arrays of the treatments were also highly regarded, but more and clearer information was needed in the ‘advice and guidance’ treatment option card. Some participants expressed that they needed more guidance and support related to the decision-making process about a specific treatment, and stated they would have liked more reflective questions on how to decide between the options. Both the patients with SAPS and healthcare practitioners found that the decision aid was easy to use and understand. However, they would have appreciated more detailed explanation and education in terms of its intended use. All healthcare practitioners said that they would have wanted a “*quality-indicator*” or “*certainty-indicator*” which could be used to grade the quality of evidence supporting each treatment. Furthermore, two healthcare practitioners expressed that they would have liked a card showing the effectiveness of psychological treatments, including behaviour-change interventions. Lastly, all participants expressed that they liked that the decision aid was delivered in a physical copy, but that they would also have liked an online version of the aid. Several components were not frequently used in the consultation; the surgery card was rarely used, and two patients stated that the ‘last option’ card was unnecessary because they would regard these options as alternative therapies. Additional examples of statements regarding the use of the decision aid can be seen in **Table 3**.

**Table 3.** Usability Statements.

| Table 3. Usability Statements |  |  |
| --- | --- | --- |
| Component | Quote | Patient or clinician |
| Information | *"I really like how there's a lot of different information, but the way it you have it structured made it much more digestible"* | Patient 2, general practice |
| Workflow | *"There's a lot going on when patients first come to us with shoulder pain, and while I try to give them the decision aid, it's hard to focus on that when we're dealing with their pain and limitations."* | General practitioner |
| Visuals | *"The visuals kept me motivated to read further; the design was just generally very eye-pleasing"* | Patient 5, PT-clinic |
|  | *"It looks really professional, so I think it is kind of cool to give it to my patients – I think it looks like we are giving them something extra, something special"* | Physiotherapist 2 |
| Sections missing | *"I was wondering why there was no graphs for the advice section, also I kind of wanted a section on natural healing"* | General practitioner |
| Content missing | *"I would have been great with a QR-code or something which would have linked to the online version"* | Physiotherapist 2 |
|  | *"I would also have liked something that could address if there was any uncertainties in terms of the treatments – like should we just completely discard surgery or is there someone who will actually benefit from it"* | General practitioner |
|  | *“Similarly, I would also have liked that would have been some information on whether the results applied to certain groups? Like is exercise only effective for certain groups or?”* | General practitioner |
| Other treatments | *“You know some of the patients definitely needs some psychological treatment, it would be nice if a future edition would have information on that”* | Physiotherapist 1 |
| Sections not used | *“Nope – I haven’t seen the disclaimer”* | Patient 18, PT-clinic |
|  | *“I haven’t had a single patient, that have used the questions that was suggested, maybe it should be highlighted a bit more”* | Physiotherapist 1 |
| Delivery | *“I think it was quite easy to use, but I was a bit unsure about what to with it all after the first time I had it delivered”* | Physiotherapist 2 |
| Language | *“Honestly, some of the writing was a bit difficult, I like the rest but especially the information not provided on the cards were a bit difficult”* | Patient 23, general practice |
|  | *“I was thinking about whether there was any version in different languages. My dad would have benefitted from this as well, but he can’t speak Danish”* | Patient 9, PT-clinic |
| Locating the relevant information | *“Easy. I think it was an easy read. It was super easy to locate what was relevant to me based on the writings and the visuals”* | Patient 14, PT-clinic |
| Comparison | *“I know I was able to just have the cards spread out, but I kind of missing on card just showing all the graphs side-by-side”* | Patient 10, general practice |
| Writing | *“I like how the information was provided in a neutral-tone, many of the websites I have seen are either very-much pro exercise or surgery”* | Patient 2, general practice |
| Trusting the information | *“I don’t know, I might have liked if there was some “patient-stories” like Trustpilot. I really like when I can mirror myself in the experience”* | Patient 13, PT-clinic |

### Consequences of using the decision aid

From the beta-testing, no unanticipated consequences (e.g., misleading content) were reported after the use of the decision aid. Several participants expressed through the interviews that the decision aid equipped them with the appropriate knowledge to decide on their treatment(s). Others expressed that they needed additional sessions with their healthcare practitioners to discuss their new insights and knowledge. Healthcare practitioners expressed that the decision aid helped them involve their patients more (i.e. shared decision-making), even in other conditions not related to shoulder pain. Neither patients nor healthcare practitioners expressed that any perception that using the decision aid delayed treatment initiation. Similarly, healthcare practitioners stated that the decision aid didn’t interfere significantly with their daily workflow and schedule. Some patients expressed that the decision aid made them more certain in their own decisions and raised their expectations of the outcome of treatment. All three healthcare practitioners stated that they also liked the decision aid because it gave them more certainty in their own management of the patient and provided them with an update on research related to treating shoulder pain.

### Degree to which the decision aid was delivered and used as intended (fidelity)

The purpose of the decision aid was not for healthcare practitioners to guide patients through the entire decision process but rather to collaborate closely with the patient to identify and discuss the most relevant treatment options. In most cases, the decision aid was used as intended. However, in some instances, the entire content of the decision aid was discussed between the patient and clinician. In a few cases the decision aid was discussed with very little information given by the clinician. Healthcare practitioners stated that their use (i.e. how much they explained about the decision and how much they discussed each component) of the decision aid varied depending on the number of patients they had to see that day and work-pressure, where the decision aid was discussed in less detail on busier days. Based on healthcare practitioners’ responses, it was estimated that using the decision aid took an average of 3-5 minutes per consultation, which was nearly the same amount of time they previously spent discussing treatment options before its introduction. In this regard, healthcare practitioners mentioned that the decision aid helped them structure their history taking with patients, especially related to the treatment decision-making process.

### Changes made from beta-testing

Several changes were made based on the feedback from beta-testing, primarily focused on adding or changing information rather than removing information (see **table 4**). Several changes suggested by participants were not implemented in the final decision aid. These included adding patient voices, psychological treatments, a card comparing all treatments side-by-side, and the costs of treatments. We did not add patient quotes for each treatment because it could introduce variability in how the treatments are perceived, potentially overshadowing the objective presentation of evidence-based information, and because a recent study has questioned the effectiveness of such narratives [51]. The inclusion of psychological treatment as an option was not implemented in the physical version because it is uncommonly used as a primary treatment for SAPS and lacks high-quality evidence. A card showing the benefits and harms of all treatments side-by-side was not added because the current layout of the decision aid allowed users to use the physical cards to compare treatments side-by-side. In our Danish primary care context, treatments rely on the tax-payer funded public healthcare system, and only to a lesser degree on individual health plans. Thus, cost information was not added. The full decision aid created based on the beta-testing can be seen in **Appendix 7**.

**Table 4.**
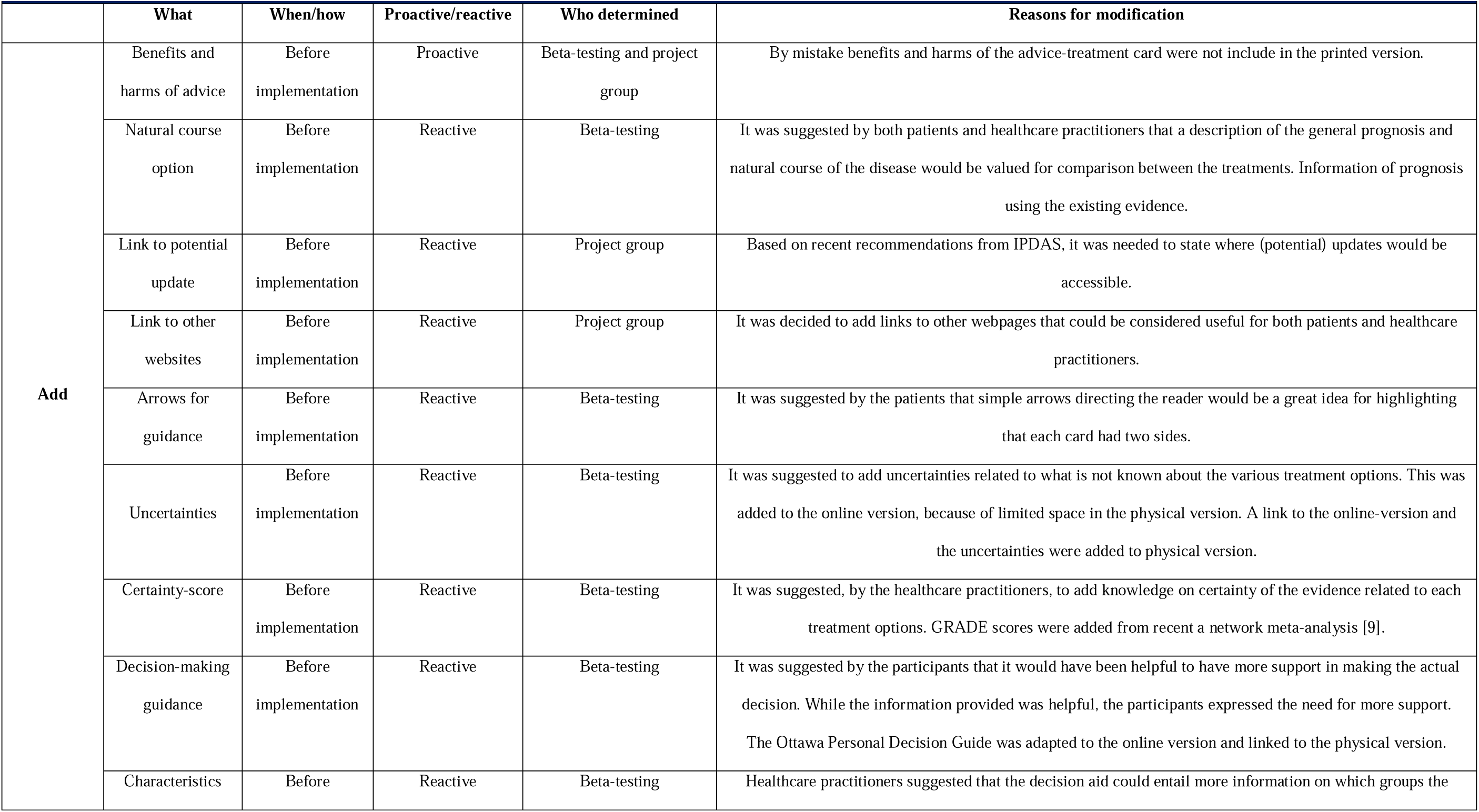
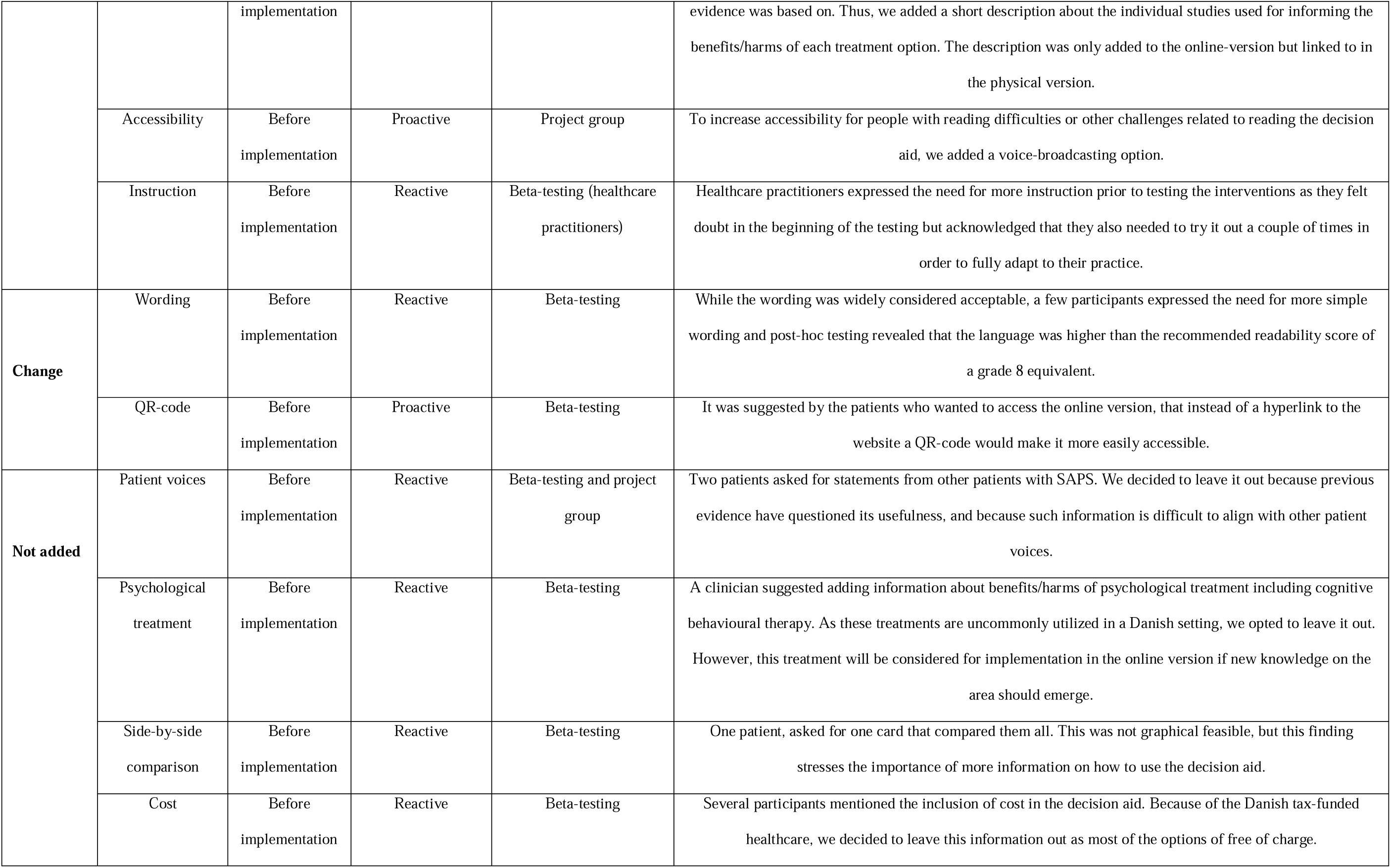
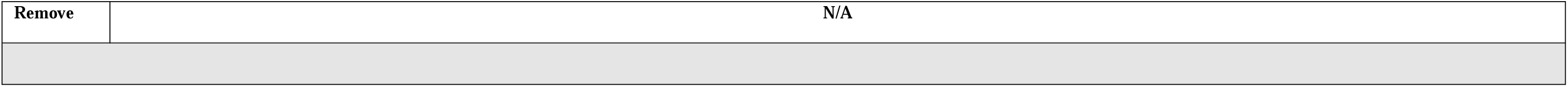
Actions taken based on the feedback from activity IV.

| Table 4. Actions taken based on the feedback from activity IV |  |  |  |  |  |
| --- | --- | --- | --- | --- | --- |
|  | What | When/how | Proactive/reactive | Who determined | Reasons for modification |
| Add | Benefits and harms of advice | Before implementation | Proactive | Beta-testing and project group | By mistake benefits and harms of the advice-treatment card were not include in the printed version. |
|  | Natural course option | Before implementation | Reactive | Beta-testing | It was suggested by both patients and healthcare practitioners that a description of the general prognosis and natural course of the disease would be valued for comparison between the treatments. Information of prognosis using the existing evidence. |
|  | Link to potential update | Before implementation | Reactive | Project group | Based on recent recommendations from IPDAS, it was needed to state where (potential) updates would be accessible. |
|  | Link to other websites | Before implementation | Reactive | Project group | It was decided to add links to other webpages that could be considered useful for both patients and healthcare practitioners. |
|  | Arrows for guidance | Before implementation | Reactive | Beta-testing | It was suggested by the patients that simple arrows directing the reader would be a great idea for highlighting that each card had two sides. |
|  | Uncertainties | Before implementation | Reactive | Beta-testing | It was suggested to add uncertainties related to what is not known about the various treatment options. This was added to the online version, because of limited space in the physical version. A link to the online-version and the uncertainties were added to physical version. |
|  | Certainty-score | Before implementation | Reactive | Beta-testing | It was suggested, by the healthcare practitioners, to add knowledge on certainty of the evidence related to each treatment options. GRADE scores were added from recent a network meta-analysis [9]. |
|  | Decision-making guidance | Before implementation | Reactive | Beta-testing | It was suggested by the participants that it would have been helpful to have more support in making the actual decision. While the information provided was helpful, the participants expressed the need for more support. The Ottawa Personal Decision Guide was adapted to the online version and linked to the physical version. |
|  | Characteristics | Before | Reactive | Beta-testing | Healthcare practitioners suggested that the decision aid could entail more information on which groups the |
|  |  | implementation |  |  | evidence was based on. Thus, we added a short description about the individual studies used for informing the benefits/harms of each treatment option. The description was only added to the online-version but linked to in the physical version. |
|  | Accessibility | Before implementation | Proactive | Project group | To increase accessibility for people with reading difficulties or other challenges related to reading the decision aid, we added a voice-broadcasting option. |
|  | Instruction | Before implementation | Reactive | Beta-testing (healthcare practitioners) | Healthcare practitioners expressed the need for more instruction prior to testing the interventions as they felt doubt in the beginning of the testing but acknowledged that they also needed to try it out a couple of times in order to fully adapt to their practice. |
| **Change** | Wording | Before implementation | Reactive | Beta-testing | While the wording was widely considered acceptable, a few participants expressed the need for more simple wording and post-hoc testing revealed that the language was higher than the recommended readability score of a grade 8 equivalent. |
|  | QR-code | Before implementation | Proactive | Beta-testing | It was suggested by the patients who wanted to access the online version, that instead of a hyperlink to the website a QR-code would make it more easily accessible. |
| **Not added** | Patient voices | Before implementation | Reactive | Beta-testing and project group | Two patients asked for statements from other patients with SAPS. We decided to leave it out because previous evidence have questioned its usefulness, and because such information is difficult to align with other patient voices. |
|  | Psychological treatment | Before implementation | Reactive | Beta-testing | A clinician suggested adding information about benefits/harms of psychological treatment including cognitive behavioural therapy. As these treatments are uncommonly utilized in a Danish setting, we opted to leave it out. However, this treatment will be considered for implementation in the online version if new knowledge on the area should emerge. |
|  | Side-by-side comparison | Before implementation | Reactive | Beta-testing | One patient, asked for one card that compared them all. This was not graphical feasible, but this finding stresses the importance of more information on how to use the decision aid. |
|  | Cost | Before implementation | Reactive | Beta-testing | Several participants mentioned the inclusion of cost in the decision aid. Because of the Danish tax-funded healthcare, we decided to leave this information out as most of the options of free of charge. |

|  |  |
| --- | --- |
| Remove | N/A |

## Discussion and Conclusion

### Discussion

In this mixed-method study, we adapted a previously tested decision aid, developed new items, and evaluate our new decision aid for care-seeking patients diagnosed with SAPS in primary care. The new decision aid was well-received by patients and healthcare practitioners during both the alpha-and beta-testing. Both groups highlighted its potential for use in clinical practice to facilitate engagement in treatment decision-making. The beta-testing results indicated that patients experienced low levels of decisional conflict and regret two weeks after being introduced to the decision aid. These findings indicate that our decision aid has the potential to enhance shared decision-making in the management of SAPS.

Within the last decade, research related to health has shifted away from a paternalistic approach to a more collaborative and patient-centered approach. Decision aids have been used to support the shift towards greater patient involvement. The development of decision aids includes strong focus on co-development with patients and healthcare practitioners to facilitate uptake in clinical practice [52,53]. A recent Cochrane review found that decision aids are likely to help patients make informed choices that are congruent with their values (compared to no use of decision aids) [54]. The review further showed that decision aids provide significant improvements in knowledge, accurate risk perception, and active engagement in decision-making [54]. Despite these positive benefits, there are very few decision aids specifically targeting musculoskeletal pain such as ours [55]. Existing aids within musculoskeletal pain have been shown to increase patient knowledge about surgical versus non-surgical management but have failed to support decisions between non-surgical options [55]. In comparison, our results suggest that providing context-specific and more information on a wider range of primary care treatment options may reduce decisional conflict.

After beta-testing, decisional conflict and regret were generally low two weeks after the introduction of the decision aid. These findings might suggest that our decision aid supports patients in dealing with treatment-related uncertainty, commonly experienced by patients with SAPS [56]. This, however, needs to be tested in larger studies comparing the decision aid to a control intervention. Importantly, healthcare practitioner feedback from the beta-testing suggested that there was a need for information on the degree of certainty for each treatment in the decision aid. To ensure this, decision aids need to be transparent and based on the best available evidence, requiring continuous updates based on new evidence of higher quality [57]. This raises attention to the need for decision aids that are easy to integrate into clinical workflows and can be continuously updated in real-time concurrently with new emerging evidence (e.g. web-based versions) [16,58–60].

Previous research has highlighted the need for considering and tailoring implementation strategies for the investigation of effectiveness in decision aids [61]. Based on the interviews after beta-testing, it became evident that both the patients and healthcare practitioners used the decision aid to a various extent. This might be caused by the introduction of many different options, which may have been overwhelming to users, making it challenging to engage with the full decision aid. Furthermore, the interview data revealed that the decision aid was easily adaptable to individual needs, potentially facilitating the integration into the workflow of clinical practice as intended. These findings add to the growing literature, emphasizing the importance of co-production and context-fit to improve the applicability of decision aids to clinical practice [62–65].

### Strengths and Limitations

One key strength of our study is the use of multiple methodological steps to develop a new context-specific decision aid for patients with SAPS. This includes both an adaptation of a thoroughly tested decision aid, and the use of established frameworks for both the adaptation and development of new items. This ensured that the decision aid was rigorously developed in collaboration with different stakeholders and using the best available evidence. Furthermore, the inclusion of different stakeholders and iterative testing, increases the usefulness and meaningfulness of the final decision aid [66,67]. Our study also demonstrates limitations. As an example, we only included participants from four primary care practices in the Region of Northern Jutland in Denmark. This limits the diversity of the included sample (both patients and healthcare practitioners), potentially leaving out key regional differences in terms of approaches to care, demographics, and needs. Based on feedback from participants, we opted to add several treatment options to the decision aid, despite limited evidence available. These additions might inadvertently promote interventions that are ineffective and thus warrants further investigation in terms of how it influences the treatment-choices compared to a control group [68]. Our study only involved feedback from three healthcare practitioners, which increases the uncertainty of the *actual* clinical usability of the decision aid. To account for potential difference in specific characteristics, approaches and preferences from the healthcare practitioners, our study warrants further research with a larger and more representative sample of healthcare practitioners. Because of these limitations, caution is needed when interpreting these findings and considering wider evaluation and implementation of the decision aid. Addressing the study’s limitations is essential before large-scale testing to enhance its effectiveness and adoption in clinical practice.

### Conclusion

The study showed that our decision aid tailored towards patients with SAPS was acceptable and aligned with the values and preferences of both patients and healthcare practitioners. Our decision aid provides information about the condition and includes evidence-based information on the benefits and risks of various treatment options, supporting patients in making decisions that are consistent with their personal values and preferences. Future large-scale trials are needed to determine the benefits of the decision aid in primary care patients with SAPS.

### Practice implications

Our study succeeded in adapting, developing and evaluation a context-specific decision aid for patients with SAPS in primary care in Denmark. Therefore, this decision shows potential in supporting SDM, reducing decisional conflicts and alleviate decisional regrets in clinical practice. Importantly, the effectiveness of the decision aid needs to be evaluated in a larger-scale study.

## Data Availability Statement

The anonymised raw data may be made available by the authors upon reasonable request.

## Acknowledgements

The project group would like to thank all the participants, who gave their time and commitment to support the research. Furthermore, the authors would like to thank the PPI group for providing inputs throughout the study. We are also very grateful to healthcare practitioners who recruited participants. Lastly, the authors would like to thank Laila Nygaard from Nygaard Grafisk Design who designed the print-based version of the decision aid and to Magnus Øberg Faurhøj for support during the creation of the online version.

## Author Contributions

**Conceptualisation:** KDL, SCB, JRZ, NEF, JLO, JLT, JS, GE, REE, MSR. **Methodology:** KDL, SCB, JRZ, NEF, JLO, JLT, JS, GE, REE, PM, VL, FD, MSR. **Software:** KDL. **Validation:** KDL, SCB, JRZ, NEF, JLO, JLT, JS, GE, REE, PM, VL, FD, MSR. **Data analysis:** KDL, SCB, MSR. **Investigation:** KDL, SCB, JRZ, PM, VL, FD, MSR. **Resources:** KDL, JLO, MSR. **Data Curation:** KDL, SCB. **Writing - Original Draft Preparation:** KDL, SCB, MSR. **Writing - Review & Editing Preparation:** JRZ, NEF, JLO, JLT, JS, GE, REE, PM, VL, FD. **Visualisation Preparation:** KDL, SCB. **Supervision:** JRZ, NEF, JLO, JLT, JS, GE, REE, MSR. **Project administration:** KDL, MSR. **Funding acquisition:** KDL, JLO, MSR.

