## Appendices 1-6 for "Development and Initial Evaluation of a Patient Decision Aid to Support Decision-Making in Care-seeking Patients with Subacromial Pain Syndrome in Primary Care"

**Appendix 1. ADAPT Checklist**

| **Checklist of questions to guide teams in adapting an intervention for a new context** | |
| --- | --- |
| **Item** | **Comment** |
| **Forming an adaptation team**  Have you involved an appropriate range of stakeholders, including those with expertise in the intervention and its evidence base, and those with knowledge of the new context?     - Is your team clear on roles, including who will make decisions on adaptations, when, and how? - Will you work with the developers of the intervention? If so, how will you manage any conflicts of interest arising from this? - How might the membership of your adaptation team need to evolve as adaptation progresses? | Yes, the adaptation team includes a multidisciplinary group of experts, including those with expertise in decision aids, musculoskeletal health, SAPS, shared decision-making, and the Danish healthcare context. KDL is the primary decision-making together with his PhD-supervisors (MSR, NEF, JLO, JLT, JS).    The team also includes clinicians and researchers from both Denmark and internationally (Australia, UK and USA), as well as patient and other stakeholder representatives, which ensures a broad perspective.    The developer of the adapted intervention (JRF) accepted to collaborate on this version.    The adaptation team expanded because we need more information to aid the development of the decision aid. This did not affect the team-roles. |
| **Assessing the rationale for intervention, and considering intervention-context fit**  • What is the problem that an intervention seeks to improve in the target population?      • Is there more than one potential evidence informed intervention? If so, are there reasons one might be more suitable than others, such as the relevance of its programme theory and change mechanisms?      • What is known about the selected intervention(s) in terms of programme theory, process, effectiveness, cost effectiveness and implementation in other contexts?                            • How robust are any claims that the intervention(s) has worked elsewhere?                      • How similar and different are original and new contexts, in terms of issues likely to affect implementation and effectiveness?                • Are there any intellectual property issues which limit use and adaptation of the intervention(s)? Planning for and undertaking adaptations             • What adaptations can you make to respond to constraints and facilitators, while maintaining consistency with intended intervention functions?            • What adaptations need to be made to intervention materials, such as manuals, to capture changes made to the intervention?        • Might interactions with aspects of the new context lead to any new unintended consequences?            • What costs and resources are needed to deliver the adapted intervention?      • Who will deliver the adapted intervention, and how will you recruit them? | 1. **What is the problem that the intervention seeks to improve?**  - The intervention seeks to support shared decision-making for patients with SAPS, where multiple non-surgical treatment options exist, and the clinical decision-making process is preference-sensitive due to the lack of a clearly superior treatment option.  2. **Is there more than one potential evidence-informed intervention?**  - Yes, other decision aids exist, including the one developed by Malliaras et al. However, the decision aid by Zadro et al. is more suitable due to its aim, focus on treatment, and inclusion of evidence-based treatment options.  3. **What is known about the selected intervention in terms of programme theory, process, effectiveness, cost-effectiveness, and implementation in other contexts?**  - Zadro et al.'s decision aid has been shown to be effective in improving knowledge. However it was not able to shown an effect in terms of treatment intention, attitudes, informed choice, and decisional conflict, which might be because of the timing of the delivery.   It was developed using IPDAS standards and was tested in patients considering surgery. Its effectiveness has been supported, though it requires adaptation to be relevant to the Danish context.   4. **How robust are any claims that the intervention has worked elsewhere?**  - The claims are robust based on online randomized controlled trials conducted in Australia, though the decision aid was primarily focused on surgical options, which limits its generalizability to Denmark, where non-surgical treatment is emphasized.      5. **How similar and different are original and new contexts?**  - The original Australian context differs from Denmark, particularly in healthcare system structure and treatment preferences. The Danish clinical guidelines emphasize exercise therapy as the first-line treatment, whereas the original decision aid by Zadro et al. focused on surgical options.      6. **Are there any intellectual property issues which limit use and adaptation?**  - There do not appear to be any intellectual property limitations. The decision aid will be translated and adapted following established protocols, with proper attribution to the original authors.  7. **What adaptations can you make to respond to constraints and facilitators while maintaining consistency with intended intervention functions?**  - The main adaptation is to include more comprehensive information about non-surgical treatment options, including exercise therapy, pharmacy, injections, and advice/education to align with Danish context.  8. **What adaptations need to be made to intervention materials?**  - The materials need to be translated to Danish using the TRAPD translation method. Additionally, content related to non-surgical treatments must be expanded using the IPDAS criteria and ODSF.  9. **Might interactions with aspects of the new context lead to unintended consequences?**  - Possible unintended consequences could arise if patients are confused by the expanded non-surgical options, but this will be mitigated by thorough testing (alpha and beta). Disruption in workflow might also be an unintended consequence, which will also be thoroughly tested.  10. **What costs and resources are needed to deliver the adapted intervention?**  - Costs will include printing or digital hosting of the decision aid, training for clinicians, and resources for testing (e.g., questionnaires and patient/clinician recruitment).  11. **Who will deliver the adapted intervention, and how will you recruit them?**  - The intervention will be delivered by clinicians involved in the management of SAPS patients (general practitioners, physiotherapists, etc.). Recruitment will be done through professional networks, clinical practices, and social media. |
| **Planning for and Undertaking Adaptations** | 1. **What adaptations can you make to respond to constraints and facilitators while maintaining consistency with intended intervention functions?**  - The main adaptation is to include more comprehensive information about non-surgical treatment options, including exercise therapy, pharmacy, injections, and advice/education to align with Danish context.  2. **What adaptations need to be made to intervention materials?**  - The materials need to be translated to Danish using the TRAPD translation method. Additionally, content related to non-surgical treatments must be expanded using the IPDAS criteria and ODSF.  3. **Might interactions with aspects of the new context lead to unintended consequences?**  - Possible unintended consequences could arise if patients are confused by the expanded non-surgical options, but this will be mitigated by thorough testing (alpha and beta). Disruption in workflow might also be an unintended consequence, which will also be thoroughly tested.  4. **What costs and resources are needed to deliver the adapted intervention?**  - Costs will include printing or digital hosting of the decision aid, training for clinicians, and resources for testing (e.g., questionnaires and patient/clinician recruitment).  5. **Who will deliver the adapted intervention, and how will you recruit them?**  - The intervention will be delivered by clinicians involved in the management of SAPS patients (general practitioners, physiotherapists, etc.). Recruitment will be done through professional networks, clinical practices, and social media. |
| **Planning for and undertaking evaluation**  • Given what is known about the intervention, and the likely transferability of previous evidence, what type and extent of re-evaluation is warranted?  • What will be the value of new information to policymakers, practitioners, and other stakeholders?  • What resources are available for re-evaluation?  • Does initial feasibility testing indicate that any further adaptations are needed?  • How will you capture responsive adaptations and decide whether the intervention remains consistent with intended functions and change mechanisms?  • How will you evaluate effectiveness, cost effectiveness, and process, if this is warranted by uncertainty about whether existing evidence will transfer? | •  **What type and extent of re-evaluation is warranted?**   - Alpha and beta testing will be carried out to evaluate the face validity, acceptability and utility, of the decision aid. Field testing will follow to assess real-life use.   •  **What will be the value of new information to policymakers, practitioners, and other stakeholders?**   - This adaptation could provide valuable insights into how decision aids for musculoskeletal conditions can be adapted and implemented in a different healthcare context, influencing national health policies on patient-centered care and shared decision-making.   •  **What resources are available for re-evaluation?**   - The project is supported by TrygFonden, Aalborg University and Danske Fysioterapeuter, with access to clinical sites for patient recruitment and data collection.   •  **Does initial feasibility testing indicate that further adaptations are needed?**   - Further adaptations will be guided by feedback from alpha and beta testing, ensuring that the decision aid is relevant and useful in the Danish context.   •  **How will you capture responsive adaptations?**   - Responsive adaptations will be captured through continuous feedback from users during testing, recorded and analyzed systematically.   •  **How will you evaluate effectiveness, cost-effectiveness, and process?**   - Effectiveness will be evaluated through patient-reported outcomes (e.g., Decisional Conflict Scale) and interviews, while cost-effectiveness could be analyzed if relevant data is available from field testing. |
| **Implementing and maintaining the intervention at scale**  • What long term partnerships and capacity will be needed for maintenance of the intervention?      • How will you monitor whether the intervention continues to be delivered, and maintains its effectiveness, over time in real world practice? | • **What long-term partnerships and capacity will be needed for maintenance?**   - Long-term partnerships with national health agencies and clinical networks in Denmark will be needed to maintain the decision aid's relevance and integrate it into standard practice.   • **How will you monitor whether the intervention continues to be delivered and maintains its effectiveness?**   - Monitoring will be conducted through periodic audits and feedback from clinicians, with adjustments made based on feedback, emerging evidence or clinical guidelines. |

**Appendix 2. The TIDieR (Template for Intervention Description and Replication) Checklist:**

Information to include when describing an intervention and the location of the information

| **No.** | **Item** | **Where located** |
| --- | --- | --- |
| **1** | **BRIEF NAME** | Development of a patient decision aid on subacromial decompression surgery and rotator cuff repair surgery: an international mixed-methods study |
| **2** | **WHY:** Describe any rationale, theory, or goal of the elements essential to the intervention. | To assist patients with subacromial pain syndrome in making informed decisions about surgery options. |
| **3** | **WHAT:**  Materials: Describe any physical or informational materials used in the intervention, including those provided to participants or used in intervention delivery or in the training of intervention providers. Provide information on where the materials can be accessed (e.g. online appendix, URL). | The decision aid contains visual aids, such as charts and graphs, to present the benefits and risks of surgery versus non-operative treatments. It includes information gathered from the latest clinical guidelines, Cochrane reviews, and patient testimonials. The materials were designed with simplicity to ensure accessibility to a wide patient demographic. |
| **4** | Procedures: Describe each of the procedures, activities, and/or processes used in the intervention, including any enabling or support activities. | A multi-step process was involved in developing the decision aid: conducting systematic reviews to gather evidence, stakeholder consultations with surgeons, physiotherapists, and patients, and iterative rounds of testing. The content was regularly revised after each round of patient feedback. |
| **5** | **WHO PROVIDED:** For each category of intervention provider (e.g. psychologist, nursing assistant), describe their expertise, background and any specific training given. | The decision aid was developed by a multidisciplinary team including orthopaedic surgeons, physiotherapists, researchers specializing in decision aid tools, and patient representatives. Their input ensured that the tool was clinically accurate and user-friendly for patients. |
| **6** | **HOW**: Describe the modes of delivery (e.g. face-to-face or by some other mechanism, such as internet or telephone) of the intervention and whether it was provided individually or in a group. | The decision aid was disseminated both digitally and in printed form for use in clinical consultations. Patients were guided through the tool by their healthcare providers to help them understand their options. Some patients used it independently outside of the clinic setting. |
| **7** | **WHERE**: Describe the type(s) of location(s) where the intervention occurred, including any necessary infrastructure or relevant features. | The tool was deployed in multiple healthcare settings, including orthopedic and physiotherapy clinics, as well as academic centers involved in the research. Recruitment for feedback involved participants from diverse clinical settings internationally, ensuring the aid was relevant to a broad audience. |
| **8** | **WHEN and HOW MUCH:** Describe the number of times the intervention was delivered and over what period of time including the number of sessions, their schedule, and their duration, intensity or dose. | Patients were introduced to the decision aid prior to consultation about their treatment options. The use of the decision aid was a one-off event, typically lasting the duration of the consultation or until the patient felt adequately informed to make a decision. |
| **9** | **TAILORING:** If the intervention was planned to be personalised, titrated or adapted, then describe what, why, when, and how. | The decision aid was developed to be applicable to various patient profiles, allowing for modifications in language based on literacy levels. It provided customized sections for different types of surgery (subacromial decompression or rotator cuff repair), as well as alternatives such as non-operative management. |
| **10** | **MODIFICATION**S: If the intervention was modified during the course of the study, describe the changes (what, why, when, and how). | Modifications to the tool were based on user feedback throughout the development process, with iterative changes to improve clarity, simplify language, and enhance the presentation of risk data. The developers also adapted the tool to include more personalized scenarios based on patient-specific factors, such as age and activity level. |
| **11** | **HOW WELL:** Planned: If intervention adherence or fidelity was assessed, describe how and by whom, and if any strategies were used to maintain or improve fidelity, describe them. | The success of the intervention was planned to be evaluated through user feedback, acceptability assessments, and measures of patient decision-making confidence. Metrics such as the rate of shared decision-making and patient satisfaction were key indicators of success. |
| **12** | Actual: If intervention adherence or fidelity was assessed, describe the extent to which the intervention was delivered as planned. | The tool was shown to be effective in improving patients’ understanding of their treatment options and increasing their engagement in decision-making processes. Most patients reported feeling more confident about their decisions and had a better grasp of the risks and benefits of surgery. Formal evaluations showed that the tool was highly acceptable to both patients and clinicians. |

**Appendix 3. TRAPD**

*The following appendix, describes the TRAPD process:*

Firstly, an English-to-Danish translation was performed by two authors (SB and KL) both with a professional background in physiotherapy and fluent in both Danish and English.

Text from the original decision aid was entered into a table structure, where each section of the original decision aid was assigned its own corresponding cell along with accompanying empty cells for the translations. The authors individually conducted their own translation, by filling out the empty cells in the table. Afterwards the two translations were fitted into one table alongside the original text.

Secondly, three new independent researchers, fluent in Danish and English, reviewed the two translations in comparison with the original text. All reviewers were blinded from knowing which author had made each translation. Each reviewer composed a translation based on the sections they preferred from the two authors or added their own suggestions to sections. The reviewers provided additional general feedback on the translation. The reviewers’ choices, suggestions and feedback were discussed among the two original translators and assembled into a second version of the translation.

Thirdly, three new people were assigned to adjudicate the second version of the translated text. The adjudication was done independently by two administrative colleagues and a physiotherapist with expertise in shoulder pain. The adjudicators provided feedback on the language and grammar, the coherence of the written content, and the comprehensibility for patients and other non-health professionals. They also provided feedback for adaptations to ensure linguistic and culturally appropriateness within the Danish context. Feedback from the adjudicators was reviewed and compiled into a third version by SB and KL.

Fourthly, to help figure out any issue that could have been missed during the previous steps, we conducted a think aloud exercise with ten patients who either lived with shoulder pain or had recovered [22]. Participants were asked to engage in a reading exercise where they expressed their thoughts and inquiries as they read through the translated decision aid. To document this the participants marked where they had any thoughts, which a researcher (SB) recorded by writing down all of their verbal contributions.

Description of each stage of the TRAPD process is summarized in **Table 1**, alongside a summary of the activities that were undertaken.

| **TRAPD (Translation, Review, Adjudication, Pre-testing, Documentation)** | | |
| --- | --- | --- |
| **Stage** | **Definition** | **Activities undertaken** |
| 1. Translate | First translation by two individuals who are fluent in both languages. | Two translators independently translated the decision aid from English to Danish. |
| 2. Review | The translated versions are sent to reviewers who are expected to select from the two translations and/or add translation options | Three reviewers independently checked both translations against the original version and complied individual suggestions for the decision aid. Feedback was integrated into a second version by the two original translators. |
| 3. Adjudication | The adjudication process wherein decisions on the final version of the decision aid are made | The second version was presented to three new people who contributed feedback before the third version was developed. |
| 4. Pretest | The translated decision aid is pilot tested on a sample that culturally represents the target population. | The third version was pilot tested through interviews on patients (n=10) who had or had recovered from shoulder pain. |
| 5. Documentation | The whole process is documented | All previous stages have been documented as seen below. |

***Table 1*** – Overview of the TRAPD process

**Appendix 4. Interview guide and acceptability questions**

**Interview guide**

| **Topic** | **Primary questions** | **Follow-up questions** |
| --- | --- | --- |
| Introduction and consent | - Presentation of project, aim and interviewer - Gather consent |  |
| What do you think about the length of the decision aid? | - Was it too long or too short? | - HCP only: Did using the decision aid require more, less, or the same amount of time as your usual patient interactions? |
| What do you think about the amount of information? | - Was there too much or too little? - Was there any specific section/card that was good/bad/balanced? |  |
| What do you think about the visual presentation? | - Was there any specific section/card that was good/bad/balanced? - What did you think about the logos, graphs, colours | - Was the format of the decision aid easy to use (e.g., font size, layout, navigation)? |
| How did you experience any bias towards specific treatments? | - Did you perceive any treatments to be favoured or biased towards a specific outcome or decision? | - Ask specifically to compared between each treatment |
| When do you think would have been the best time to receive/deliver the decision aid | - Would you have preferred sooner or later? - How did it fit into your daily life (or workflow) | - Do you think we could have delivered the decision aid any differently? Time, setting |
| How much and which components were used? | - How well did the decision aid help you understand your treatment options? - Were there any parts of the decision aid that were unclear or difficult to understand? | - If so, which parts? - Did you feel that the information was relevant to your specific condition and treatment options? - HCP only: Did you feel adequately prepared or trained to use the decision aid with patients? - Has the use of the decision aid led you to change any aspect of your clinical practice or communication with patients? - Were there any unexpected outcomes (positive or negative) from using the decision aid? |
| Decision-making | - How much did the decision aid help you feel more confident in making a decision? - Did it address your main concerns or questions about treatment? | - Did you feel more involved in your treatment decision after using the decision aid? |
| Why do you think it worked/not worked? | - Which components do you think helped in reaching X outcome? |  |

**Acceptability**

**Online Health Professional Acceptability Questionnaire**

| **In general:** | **Strongly agree** | **Somewhat agree** | **Neutral** | **Somewhat disagree** | **Strongly disagree** |
| --- | --- | --- | --- | --- | --- |
| It will be easy for me to use | 5 | **4** | **3** | 2 | 1 |
| It is easy for me to understand | 5 | 4 | 3 | 2 | 1 |
| It will be easy for me to experiment with using it before making a final decision to adopt it | 5 | 4 | 3 | 2 | 1 |
| The results of using the decision aid will be easy to see | 5 | 4 | 3 | 2 | 1 |
| This decision aid is better than how I usually go about helping patients decide about their shoulder pain treatment | 5 | 4 | 3 | 2 | 1 |
| This decision aid is compatible with the way I think shoulder pain should be managed | 5 | 4 | 3 | 2 | 1 |
| Compared with my usual approach, this decision aid will result in my patients making more informed decisions | 5 | 4 | 3 | 2 | 1 |
| Using this decision aid will save me time | 5 | 4 | 3 | 2 | 1 |
| This decision aid is a reliable method of helping patients make decisions about shoulder pain treatment | 5 | 4 | 3 | 2 | 1 |
| Pieces or components of the decision aid can be used by themselves | 5 | 4 | 3 | 2 | 1 |
| This type of decision aid is suitable for helping patients make value laden choices | 5 | 4 | 3 | 2 | 1 |
| This decision aid complements my usual approach | 5 | 4 | 3 | 2 | 1 |
| Using this decision aid does not involve making major changes to the way I usually do things | 5 | 4 | 3 | 2 | 1 |
| There is a high probability that using this decision aid may cause/result in more benefit than harm | 5 | 4 | 3 | 2 | 1 |

**Online Patient Acceptability Questionnaire**

| Which treatments should I have | Poor | Fair | Good | Excellent |
| --- | --- | --- | --- | --- |
| Who should read this decision aid | Poor | Fair | Good | Excellent |
| What are the treatment options covered in this decision aid | Poor | Fair | Good | Excellent |
| Comparing benefits between treatments? | Poor | Fair | Good | Excellent |
| Comparing harms between treatments | Poor | Fair | Good | Excellent |
| Summary of benefits, harms and practical issues | Poor | Fair | Good | Excellent |
| Questions to consider when talking with your health professional | Poor | Fair | Good | Excellent |

**Appendix 5. Prototype decision aid (Danish)**

Removed due to lack of valid translation.

**Appendix 6. Decision aid (pre beta-testing) (Danish)**

Removed due to lack of valid translation.
